# Effects of kinesthetic cues supported by physiotherapist during a motor training intervention with virtual reality-based games on functioning in people with Parkinson’s disease: A prospective, single-blinded, parallel-group, randomized clinical trial

**DOI:** 10.1101/2023.08.30.23294862

**Authors:** Pamela Yuki Igarasi Barbosa, Amarilis Falconi, Matheus D’Alencar, Erika Okamoto, Maria Elisa Pimentel Piemonte

## Abstract

**Background:** It’s been suggested that kinesthetic cues (KC) could be a useful tool in helping individuals improve their motor learning process by facilitating muscle activation. However, while virtual reality-based games (VRG) are becoming more popular as a intervention tool, the potential effects of KC on this kind of intervention have yet to be investigated in people with Parkinson’s disease (PwPD). Therefore, this study aimed to compare the effects of motor intervention using VRG coupled or not with KC provided by a physiotherapist (PT) during training on the functioning of PwPD.

**Methods:** Thirty-eight PwPD in 1-3 Hoehn and Yahr (HY) stage were randomized into two groups: (1) VRG with KC Group (KCG), where KC was provided by manual assistance of a PT, and (2) VRG with NO KC group (NKCG), where no PT manual assistance was provided during the training. Both groups received 8 individual sessions with 50 minutes, twice a week, for 4 weeks: 10 minutes for warm-up and 40 minutes to play 4 games from *XBOX 360* with *Kinect®* system. Outcomes were evaluated at three time points: (1) before training (BT), (2) 1 week after training (AT), and 8 weeks after that as follow-up (FU). To reach a comprehensive evaluation of functioning, several outcomes were adopted and categorized according to International Classification of Functioning Disability and Health (ICF): Geriatric Depression Scale (GDS); Montreal Cognitive Assessment (MOCA); Unified Parkinson Disease Rating Scale (UPDRS) - section 3 (UPDRS-III); and Rapid Turns Test (RTT) used to assess motor and non-motor alteration into Function domain; Balance Evaluation Systems Test (BESTest); Falls Efficacy Scale International (FES-I); Thirty-Second Walk Test (30sWT); Six-Minute Walk Test (6mWT); 5 Times Sit to Stand Test; and UPDRS section II (UPDRS-II) used to assess motor performance into Activity domain, and finally, the Parkinson Disease Questionnaire (PDQ-39) to assess quality of life into Participation domain.

**Results:** ANOVA for repeated measures showed a significant effect for evaluation-time factor only (p-value<.001), (no effect for group or evaluation-time X group interaction) for all measures of three ICF domains, excluding GDS and 6mWT. The Tukey post-hoc test confirmed significant improvements in AT that remained at FU. Conclusions: A motor intervention using VRG can improve the functioning in terms of Function, Activity, and Participation of PwPD, regardless of the KC provided by PT during the training.

## 1. Introduction

Parkinson’s Disease (PD) is the most common neurodegenerative disorder in the world after Alzheimer’s Disease [1]. This disorder is characterized by progressive loss of dopaminergic neurons in the substantia nigra pars compacta [2,3] and, most recently, associated with abnormal α-synuclein aggregation in the nervous system [4–6] that provokes typical non-motor and motor features harming functioning, independence and quality of life in individuals affected by the disease [3,7].

World Health Organization (WHO) conceptualizes functioning as the dynamic interaction between an individual’s health state and environmental and personal factors [8]. According to the International Classification of Functioning, Disability, and Health (ICF), functioning is divided into three domains: (1) Body functions and structures; (2) Activity; and (3) Participation [9]. The use of ICF is recommended by WHO [9], the European Guideline for PD [10] and the American Physical Therapy Association [11] to improve communication between healthcare professionals, researchers, and social policymakers.

Motor and non-motor dysfunctions caused by PD result in different limitations of activity, i.e., difficulties in executing daily life activities (DLA), and consequently cause participation restrictions, i.e., problems when involved in life situations [12]. Thus, the improvement and/or maintenance of functioning with greater independence, autonomy, and quality of life is the mean goal in rehabilitation. Moreover, PD is an incurable and progressive disease, therefore the search for new low-cost approaches that benefit long-term adhesion is fundamental for better results.

In the field of physiotherapy, the use of virtual reality-based games (VRG) has been proposed as a new therapeutic alternative in neurorehabilitation. The offer of visual and auditory stimuli that provide feedback about performance and increase motivation can be highlighted as a powerful advantage [13].

In PD, the therapeutic use of VRG is applicable, efficient, and safe [14]. Evidence indicates positive short-term effects of exercises with VRG, which are similar to the results of conventional physiotherapy treatment [13,15]. Considering this potential to stimulate motor control and mental aspects such as cognition and humor [16], VRG training can benefit the functioning of people with PD (PwPD).

Despite the positive evidence on VRG for PD rehabilitation and its growing use in clinical practice, to the best of our knowledge, no physiotherapy guideline has yet been established to offer directions on the proper application to improve motor function in PwPD [13]. Among these needed directions, the ideal level of physiotherapist (PT) manual assistance during the training is one of the most important of them. VRG systems provide high real-time sensory feedback but are limited to visual and auditory information [13,14]. The kinesthetic cues (KC) provided by PT manual assistance during the training could offer additional information on body position and movement, which are essential for controlling movement and balance [17]. KC may also facilitate muscle activation, improving training gains [18]. Multisensory stimulation may facilitate motor learning and plasticity, activating multiple cortical areas [19,20]. Then, it is plausible to suppose that combining KC provided by PT with visual and auditory feedback provided by the VRG system may enhance motor performance during the training and, consequently, improve rehabilitation results.

To test this hypothesis, i.e., KC can increase the VRG training gains, we conducted a prospective, single-blinded, parallel-group, randomized clinical trial comparing the effects of intervention using VRG with and without KC provided by PT manual assistance during the training to improve the functioning in terms of motor and non-motor function, activity performance and participation level of PwPD.

We believe that the results of this study may help develop guidelines for using VRG in the rehabilitation of PwPD showing whether the manual PT physical guidance during this training may or may not improve intervention results. The findings can also contribute to a better understanding of the benefit of multisensory stimulation during motor training.

## 2. Materials and Methods

### 2.1. Design

A prospective, single-blinded, parallel-group, randomized clinical trial was conducted in Sao Paulo, Brazil. Thirty-eight people diagnosed with idiopathic PD were recruited and randomly allocated into two groups: (1) VRG with KC Group (KCG) or (2) VRG with NO KC group (NKCG). Outcome measures were applied at three time points before training (BT), 1 week after training (AT), and 8 weeks after the end of the intervention, the last one is considered as a follow-up measure (FU). This study was conducted in accordance with the Declaration of Helsinki and approved by the Research Ethics Committee of the Faculty of Medicine of the University of Sao Paulo (protocol code 79419517,4,0000,0065, approval date 21 February 2018; is registered on ClinicalTrials.gov with trial registration number NCT04717271 (30 November 2020) and in agreement with CONSORT guidelines for developing randomized trials [18].

### 2.2. Participants

The subjects were recruited at Parkinson Brazil Association, a specialized reference care center for people with Parkinson’s Disease in Sao Paulo. The study was conducted in the Department of Physical Therapy, Speech Therapy, and Occupational Therapy of the Faculty of Medicine of the University of Sao Paulo, Brazil. The assessment and intervention were performed from December 2020 to April 2021.

Most participants were invited in person, and a few were contacted by phone by a PT who was not involved in the intervention. The individuals were selected according to the following eligibility criteria established for recruitment: aged between 50 and 75 years old; diagnosis of idiopathic PD according to the London Brain Bank criteria for Neurodegenerative Diseases [2]; in stages 1 to 3 of the Hoehn and Yahr Staging Scale (HY); and be in regular use of their medication treatment.

The exclusion criteria were: the presence of severe cognitive impairment with a scored ≤ 20 on the Montreal Cognitive Assessment (MOCA); signs of depression defined as ≥ 6 on the Geriatric Depression Scale (GDS-15); having others conditions, for example, visual; cardiorespiratory and/or musculoskeletal impairment that could enable or limited the safety performance in training; participating in another rehabilitation treatment and who did not agree to sign the consent term.

### 2.3. Randomization

Participants were randomized by a senior researcher to their groups after the baseline visit and before the first day of training. A simple randomization method was used to allocate participants in each group. Each participant drew one sealed envelope, which was opened individually by the PT, who performed the intervention only when each participant started the training. During assessments, participants were instructed not to reveal any details about their training to preserve the blinded examiner.

### 2.4. Outcome measure

The three assessment points (BT, AT, and FU) were conducted by a blinded and experienced PT. To assess the functioning level of PwPD, several recommended tools were categorized according to ICF in three domains: Function, Activity, and Participation [21–23]. In order to reach a comprehensive evaluation of functioning, the eleven tools were similarly treated.

The first domain, Function, included tools to assess motor, cognitive and emotional function. Then, the GDS-15, which is composed of 15 questions, was adopted to assess signs of depression when the final score is equal to or higher than 6 [24]; MOCA is a brief cognitive screening tool used to assess seven cognitive domains (visuospatial and executive Function; naming; memory; attention; language; abstraction; orientation) with a total possible score of 30 points; a score of 26 or above is considered normal; a score between 21 and 26 detects mild cognitive impairment (MCI); while a score less than 20 points indicate severe cognitive impairment [25]; the Unified Parkinson Disease Rating Scale (UPDRS) - section 3 (UPDRS-III) to assess the severity of motor symptoms through 14 items that analyze PwPD motor performance with 0 to 4 points per items wherein 0 is considered normal whereas 4 is the most severe motor impairment [26].

In the second domain, Activity, there were several tools to assess the performance of several tasks. The Balance Evaluation Systems Test (BESTest) to assess the balance state in 36 items, grouped into 6 systems: “Biomechanical Constraints”, “Stability Limits/Verticality”, “Anticipatory Postural Adjustments”, “Postural Responses”, “Sensory Orientation”, and “Stability in Gait”, each item is scored on a 4-level, 0 (worst performance) to 3 (best performance) [27]. The Falls Efficacy Scale International (FES-I) assesses self-confidence in balance with 16 questions about the concern of falling while performing some activities, grading scores of 1 to 4, the total score can range from 16 (absence of concern) to 64 (extreme concern) [28].

Also, the Thirty-Second Walk Test (30sWT) to assess the short-distance gait performance, where the participant walks for thirty seconds under single (ST), and dual task condition (DT) while the distance is registered by the examiner [29,30]. Whereas the long-distance gait performance was assessed with the Six-Minute Walk Test (6mWT), where the participant walks thirty meters as fast as they can for 6 minutes, at the end the distance walked by the subject is measured [31].

Addiotionally, the Five Times Sit to Stand Test (FTSTS) was used to assess the performance to transfer from seat position to stand position, five times as fast as they can without using their hands while the task is timed [32,33]. The Rapid Turns Test (RTT) to assess the ability to execute rapid full turns (360º; 3 into each direction) and the presence of freezing of gait (FOG) [22,34]. Finally, the UPDRS – section II to assess the motor performance in DLA with 13 items, the score results from the self-report of the PwPD based on their last 15 days, ranging from 0 (normal) to 4 (severe impairment) [26].

The last domain, Participation, included the Parkinson’s Disease Questionnaire (PDQ-39) to assess the quality of life into eight dimensions of health (mobility, ADL, emotion, stigma, social, cognition, communication, and body pain) that PwPD report as adversely affected by the disease, then it is scored on a scale of 0 to 100, lower scores indicate better-perceived health status [35].

### 2.5. Intervention

The game system used was the Microsoft *Kinect®* for *XBOX 360*. It has a movement detector with infrared signal sensors that captures changes in the player’s direction; velocity; and acceleration, making it possible to control the player’s avatar directly with their real full-body movement. Its use has already proven safe and feasible in PwPD [36].

Participants performed 8 individual training sessions (50 minutes, twice a week, for 4 weeks) performed by a trained PT. In the first 10 minutes of each session, participants in both groups performed general exercises, including lower limb stretching; active and resistive exercises for the trunk, neck, and upper limbs, as a form of warm-up and preparation for the second part of the training, supervised by a PT (Supplemental Data File 1).

The following 40 minutes of the treatment consisted of VRG training conducted by PT, including 4 games selected from *XBOX 360* with *Kinect®* system: (1) Light Race which demands fast and large steps; (2) Stack Em up, which demands multidirectional postural responses under multitask; (3) Wall Breaker which demands quick cross movements of arms and legs as dodge the body down and to the side; and (4) Run the World which demands stationary gait with high knees. Except for the last one, that was played for five consecutive minutes, all games had three different difficulty levels (easy, medium, and hard), requiring a score of at least 200 points to pass from the easy level to the medium and 300 points from the medium to the hard level. (Supplemental Data File 2). The games were selected by four PTs with an extensive experience in the intervention using VRG in PwPD, according to their motor demand, focusing on balance and gait.

In order to facilitate the movement detection by *XBOX 360* with *Kinect®* system, a visual mark, an “X,” was placed on the floor 3 meters away from the movement detector. This was the ideal distance so the infrared sensors would capture the participant’s full-bodied movements. Furthermore, the visual mark helped participants with spatial awareness information, preventing them from leaving the sensor area, impairing their score, or even pausing the game automatically.

Finally, the PT position behind the participant during the training was crucial to guarantee proper movement detection by Kinect®.

Initially, instructions were provided about the goals, rules, and strategies to control the avatar in the game so that the participant could be familiarized with the game’s system. Afterward, the PT made a demonstration of how to play. Then, the subject was invited to begin playing the games by themselves. Motivational verbal cues were systematically similar for both groups (predefined motivational phrases at specific times in the game, for example: “go ahead,” “continue, you are doing well”), and the PT offered safety during the whole intervention.

#### 2.5.1. Virtual reality-based games with kinesthetic cues group (KCG)

In addition to visual and auditory feedback provided by *XBOX 360* with *Kinect®* system, on the first gameplay attempt, the PT stood behind the participant and provided KC by positioning her hands either on the trunk or shoulders, or even on the elbows or both iliac crests, according to each game, remaining a manual contact enough to guide the body movement without risk of skin injury or pain. The KC aimed to facilitate proper motor responses regarding timing, speed, direction, and amplitudes. The intensity and duration of KC were adapted according to motor game demand. More detailed information about the KC is shown in Table 1. The PT adapted the duration and intensity of KC from the first to the last session according to the participant’s performance improvements.

**Table 1.** KC description.

| Game | Where | How | When |
| --- | --- | --- | --- |
| Light Race | Stimuli apply on both iliac crests. | Helping to move the body in the correct direction and to put weight on the support member while the other performed the step and gave the proper impulse during the jump. | Every time one piece lighted and when two pieces lighted simultaneously, which required a jump to be able to step on both pieces at the same time. |
| Stack Em up | Stimuli apply either on the trunk or elbows, or even on the iliac crests. | Improving the anteroposterior and lateral trunk alignment. On elbows were applied to improve the inclination or stabilization of the board. The iliac crests received pressure in order to maintain balance. | The stimuli on the trunk were always given in the presence of misalignment, on elbows to balance the cubes and at the time to lean to the right or left in order to deposit the cubes safely in the correct place, on the iliac crests during the unipodal support. |
| Wall Breaker | Stimuli on shoulder or iliac crests | The KC on the shoulder was intended to direct and increase the speed of movement, a more smooth touch was applied on the iliac crests to indicate which member should perform the kick. | During the punch and the kick. |
| Run the World | Stimuli on the shoulder | KC on shoulder to improve dissociation. | At the beginning of the game and every time the dissociation started to decrease. |

#### 2.5.2. Virtual reality-based games with NO kinesthetic cues group (NKCG)

Participants performed the game with only visual and auditory feedback from the *XBOX 360* with *Kinect®* system. No KC was offered on the two attempts. PT’s participation was restricted to ensure the participant’s safety.

### 2.6. Data analysis

The sample size calculation was based on the BESTest. Based on a previous study similar to the present study in terms of volume, duration, and features of intervention and studied population, there was a difference of 2.83 points between the baseline and after training. The sample size calculation showed that 40 participants (20 in each group) would be enough for a power greater than 90% (P=0.05).

Demographic and clinical characteristics of the KCG and the NKCG participants were compared using the unpaired *t*-test or Mann-Whitney U test, according to the variable characteristics. The Kolmogorov Smirnov test was used for the distribution of variables–Smirnov normality test.

In order to analyze the game’s performance improvements, the final scores provided by *XBOX 360* with *Kinect®* system, four two-way ANOVA with repeated measures (RM-ANOVA) were used, one for each game, having as the main factor the group (KCG and NKCG) and as repeated measure the training session (8 sessions).

Following, we conducted eleven RM-ANOVAs to analyze the results from independent tools used to assess the functioning according to the ICF framework. The factors were group (KCG, NKCG) and assessment time (BT, AT, and FU), with the latter as repeated measures. A post-hoc Tukey–Kramer test was performed for effects that reached statistical significance. The intention-to-treat analysis was applied with the simple imputation of missing variables by the last observation carried forward. Differences were considered significant when *P* <0.05. The statistical analysis was performed using Statistica Version 13 (TIBCO Software Inc. USA).

## 3. Results

### 3.1. Participants

A total of 60 participants were contacted, and among them, 43 were screened. Thirty-eight PwPD (30 men and 8 women) with a mean age of 63.78 years (SD 7.58) were randomized and started the intervention. Participants were in stage I, II, or III according to the HY. During the intervention, 6 subjects were lost (Figure 1) but were analyzed by intention to treat. Demographic and clinical characteristics of the participants at baseline are presented in Table 2. There were no significant differences between groups. No adverse events were reported in both groups.

**Table 2.** Demographic and clinical characteristics of the participants at baseline.

|  | KCG (N=19) | NKCG (N=19) | p-value |
| --- | --- | --- | --- |
|  | Median (SD) | Median (SD) |  |
| Age (years) | 65.35 (5,04) | 62.21 (9,36) | 0.20 |
| Mass (kg) | 72.57 (11.09) | 72.25 (11.89) | 0.93 |
| Height (cm) | 167.26 (7.83) | 168.21 (10.34) | 0.75 |
| Years of study | 14.05 (3.61) | 13.63 (5.61) | 0.78 |
| MOCA | 25.21 (2.46) | 24.57 (2.87) | 0.72 |
| GDS | 3.78 (3.47) | 4.33 (2.49) | 0.58 |
| UPDRS total score (session II + III) | 26.63 (10.64) | 22.26 (11.49) | 0.23 |
| Gender (Relation Men/Women) | 15/4 | 15/4 |  |

KCG: virtual reality-based games with kinesthetic cues group (KCG); NKCG: virtual reality-based games with NO kinesthetic cues group (KCG); SD: Standard Deviation; MOCA: Montreal Cognitive Assessment; GDS: Geriatric Depression Scale; UPDRS: Unified Parkinson's Disease Scale.

**Figure 1.**
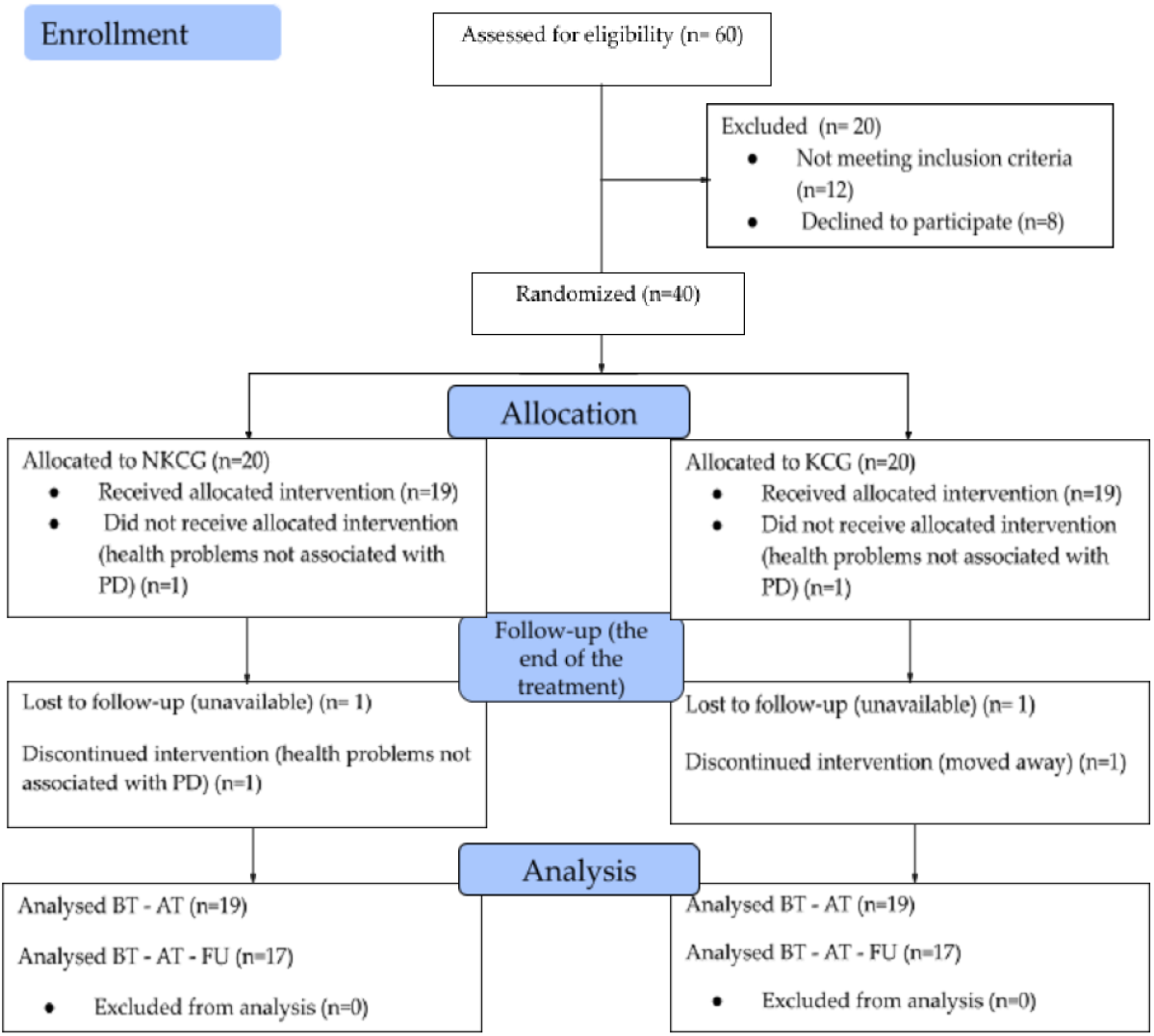
The CONSORT flowchart. NKCG: virtual reality-based games with NO kinesthetic cues group (KCG); KCG: virtual reality-based games with kinesthetic cues group (KCG); PD: Parkinson’s disease; BT: before training; AT: after training; FU: follow-up.

### 3.2. Games improvement

The RM-ANOVA for final scores in the games, showed a significant effect of assessment, with no significant effect for group or factor interaction for Light Race (F_7,25_ = 0,33; p-value < .009; Effect size = .95), Stack Em up (F_7,24_ = 1.09; p-value < .003; Effect size = .95), and Run the World (F_7,25_ =1.73; p<.001; Effect size = .95) games. The post-hoc test confirmed significant improvements between the first and the last session (S1XS8) for all games (p-value<.001).

In contrast, for Stack Em up, there was a statistically significant interaction between group factor and session (F_7,25_ = 2.60; p-value<.001; Effect size = .95). The post-hoc test confirmed that KCG reached higher scores than NKCG (p-value<.001).

Summarizing, the KC promoted an increased improvement in one of the four played games (Stack Em Up).

### 3.3. Intervention effect

Eleven recommended tools for evaluating PwPD were organized according to the ICF framework to measure possible changes in the functioning level of the participants before and after the intervention. These tools were treated similarly in order to achieve a comprehensive evaluation of functioning.

#### 3.3.1. ICF Function domain

Concerning motor function, the RM-ANOVA for UPDRS III used to assess the severity of motor symptoms showed a significant effect of assessment, with no significant effect for group or interactions between factors in the total score in (F_2,72_ = 6.60; p-value < .0002; Effect-size = .90). The post-hoc test confirmed a significant improvement after training for all of them (p-value < .0002), which remained at the follow-up (p-value < .03), regardless of group (Table 3).

**Table 3.**
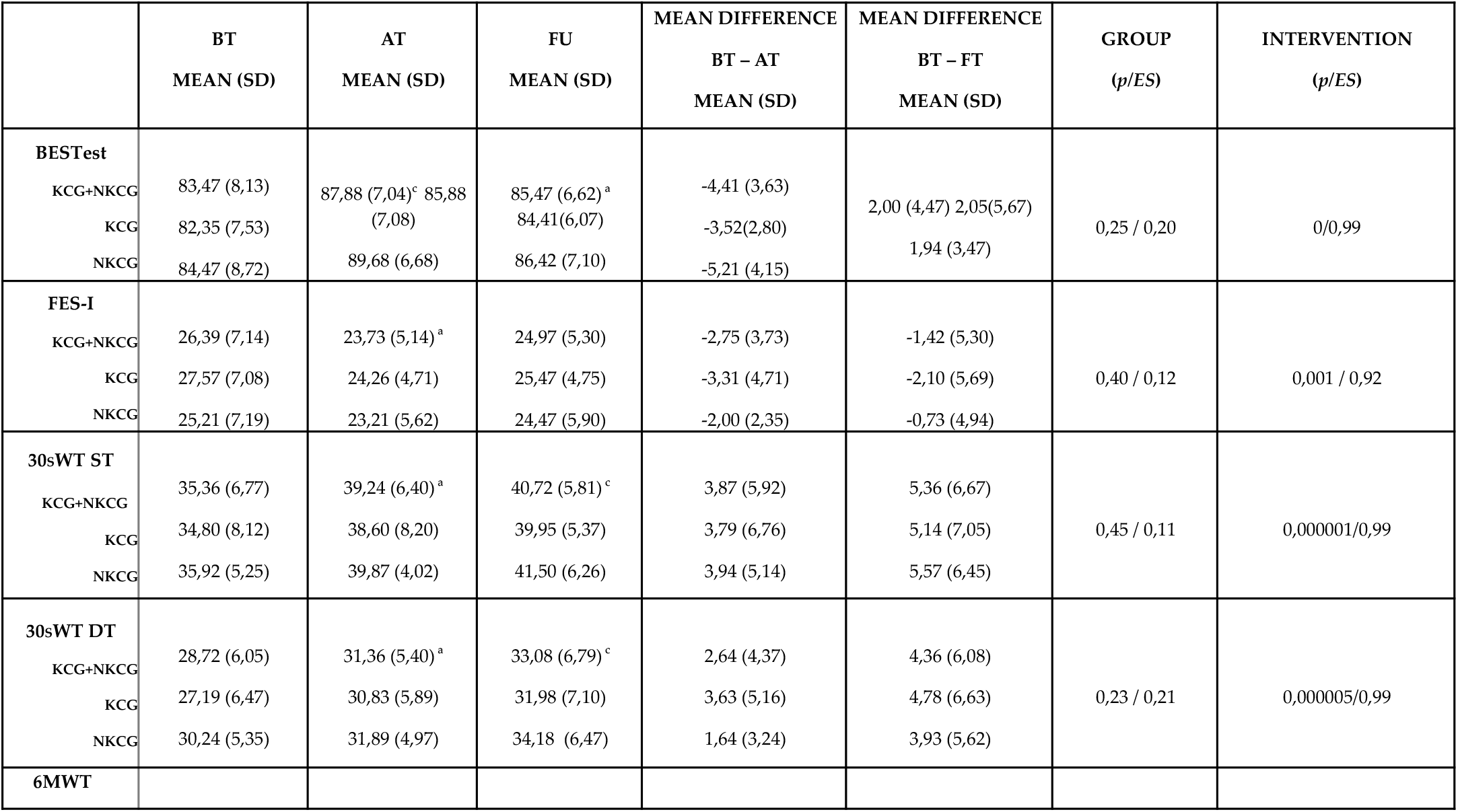
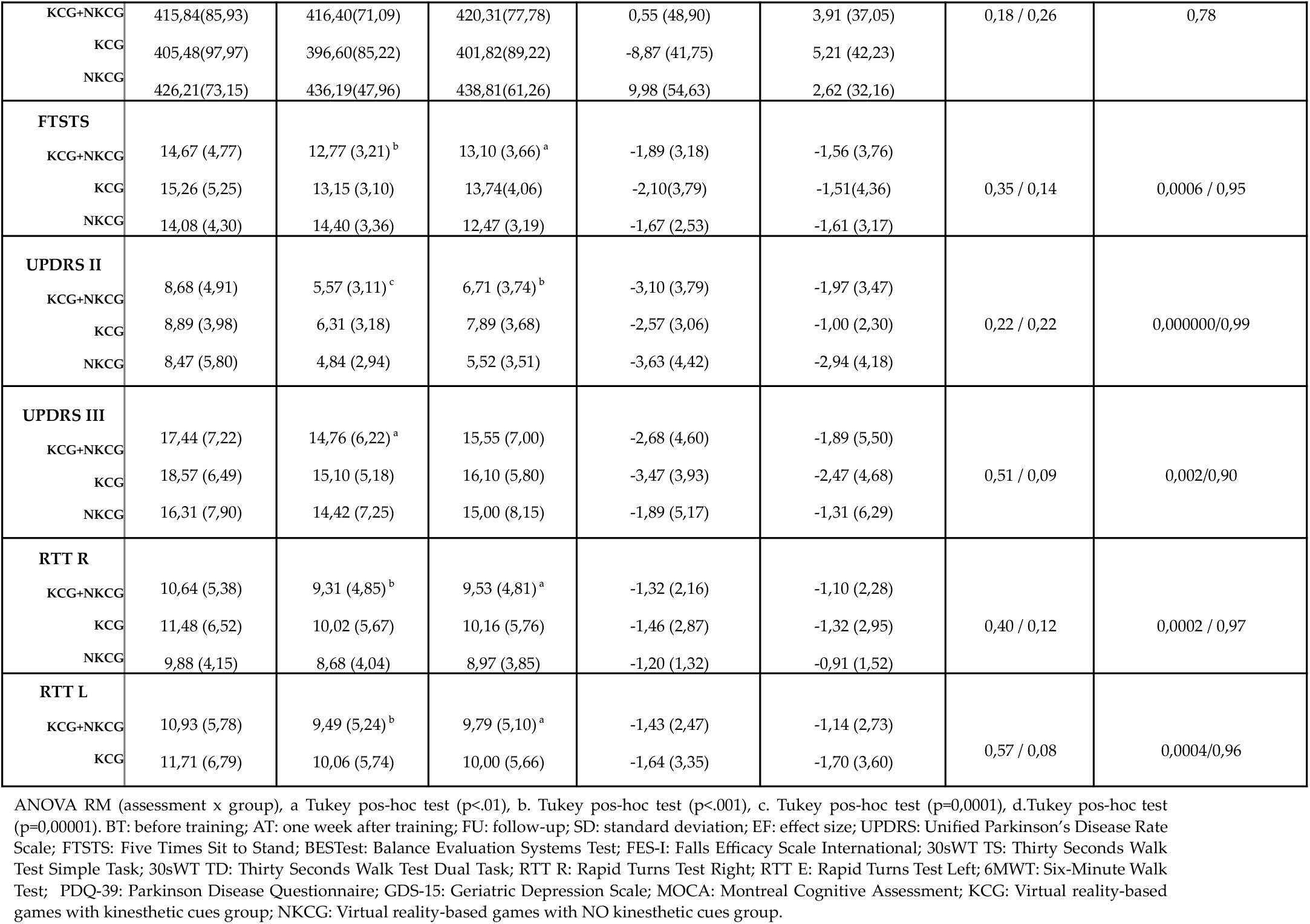
Mean (SD) for each measure in each group before and after intervention and at follow-up.

The same was observed for cognitive Function assessed by MOCA, the RM-ANOVA showed a significant effect of assessment (F_2,72_ = 8.95; p-value < .0003; Effect-size = .96), with no significant effect for group or interactions between factors. The post-hoc test confirmed a significant improvement after training (p-value < .0008), which remained at the follow-up (p-value < .002), regardless of group (Table 3).

In contrast, for mental Function assessment by GDS-15, RM-ANOVA showed no significant effect of assessment, group, or interactions between them for

Summarizing, both interventions promoted improvements in motor and cognitive function without advantages for the group that received f KC during the training. No improvement in mental Function was observed after both interventions.

#### 3.3.2. ICF Activity domain

The RM-ANOVA for total scores obtained with BESTest used to assess the balance showed a significant effect of evaluation time (F_2,68_ = 19.91; p-value < .0001; Effect-size = .999), with no significant effect for group or interaction between factors. The post-hoc Tukey test confirmed significant improvements after training (p-value < .0001) that was maintained at follow-up (p-value < .014), regardless of group (Table 3).

Congruently, the RM-ANOVA for FES-I scores used to assess the self-balance confidence: showed a significant effect for evaluation time (F_2,72_ = 7.25; p-value < .001; Effect-size = .926), with no significant effect for group or interaction between factors. The post-hoc Tukey test confirmed that there were improvements after training (p-value < .0009), regardless of group (Table 3).

The same could be observed for the RTT, which assess the motor performance during the turn: the RM-ANOVA also showed a significant effect of evaluation for total time in RTT to the right (F_2,68_ = 9.23; p-value < .0002; Effect-size = .971) as much as to the left (F_2,68_ = 8.53; p-value < .0004; Effect-size = .960) with no significant effect for group or interaction between factors. The post-hoc Tukey test confirmed an improvement after training (p-value < .001) that was maintained at follow-up (p-value < .01) for both sides, regardless of group (Table 3).

Concerning the gait performance for short distances, RM-ANOVA showed a significant effect for evaluation for maximal distance walked in the 30sWT under ST (F_2,72_ = 16.57; p-value < .000001; Effect-size = .999), and DT (F_2,72_ = 14,56; p-value < .000005; Effect-size = .998), with no significant effect for group or interaction between factors. The post-hoc Tukey test confirmed an improvement after training for all conditions (p-value < .001) that was maintained at follow-up (p-value < .001), regardless of group (Table 3).

However, concerning the gait performance for long-distance, the RM-ANOVA showed no significant effect for evaluation, group, or interaction between them for maximal distance walked in the 6mWT.

The RM-ANOVA for FTSTS used to assess motor performance during postural transfer showed a significant effect for evaluation (F_2,72_ = 8.19; p-value < .0006; Effect-size = .953), with no significant effect for group or interaction between factors. The post-hoc Tukey test confirmed an improvement after training (p-value < .0009) that was maintained at follow-up (p-value < .007), regardless of group (Table 3).

Finally, RM-ANOVA showed a significant effect for evaluation in total score in UPDRS – Section II, used to assess the impact of motor symptoms on DLA (F_2,72_ = 19.07; p-value < .000000; Effect-size = .999) with no significant effect for group or interaction between factors. The post-hoc Tukey test confirmed an improvement after training (p-value < .0001) that was maintained at follow-up (p-value < .0007), regardless of group (Table 3).

Summarizing, both trainings promoted improved balance, short distance gait under different conditions, postural transfer, and, most importantly, independence in DLA, with no advantage for the group that received KC during the training.

#### 3.3.3. ICF Participation domain

Finally, for PDQ-39 used to assess the quality-of-life associated with PD, the RM-ANOVA showed a significant effect of assessment F_2,68_ = 17.57; p-value < .000001; Effect-size = .999), with no significant effect for groups or interactions between factors. The post-hoc test confirmed a significant improvement after training (p-value < .002), regardless of group (Table 3).

## 4. Discussion

This study is the first to investigate the effects of KC offered by PT manual assistance during the training based on VRG on the functioning of PwPD. The results showed that the addition of KC did not provide any superior results to improve the functioning according to ICF, contradicting our primary hypothesis. However, both groups showed significant improvements in all tools related to Function, Activity, and Participation in ICF domains, excluding the long-distance gait and depression.

It is generally accepted that augmented feedback, provided by a human expert or a technical apparatus, effectively enhances motor learning [37]. Augmented feedback, also known as extrinsic feedback, is defined as information that cannot be elaborated without an external source; thus, it is provided by a trainer or a system [38,39]. VRG offers real-time auditory and visual feedback, which can be considered one of the most important advantages in rehabilitation use [13]. However, most VRGs are not able to offer kinesthetic feedback or guidance. The KC can physically guide the subject through the ideal motion to reach the best performance [40]. In the present study, the KC provided by PT during the training had as its main purpose to teach the proper movement parameters such as timing, direction, and range to guide the subject toward, and not necessarily through, the desired motion.

Although adding the KC did not result in higher functioning improvements compared to identical training without it, this kind of cue increased the gains for one game, Stack Em Up. Two factors may have contributed to this result: the game’s and KC’s features. Concerning the game’s features, this game requires a high level of postural control as participants had to be able to balance on just one leg while also completing multiple cognitive tasks. The addition of KC provided by PT could have facilitated learning for this game, which demanded a more complex automatic motor control under attention division. Actually, PwPD has higher difficulty performing multiple tasks simultaneously than healthy individuals due to limited attentional resources, defective central executive Function, and less automaticity in performing motor tasks [41]. The positive effect of multisensory feedback on the motor learning process increases as task complexity increases [42,43]. Supposedly, for other games, the visual and auditory feedback provided by the system was enough to promote significant learning. It is well known that visual and auditory cues can minimize the disruption in internal cues associated with dopamine depletion [44]. Several studies have shown that visual and auditory cues can improve motor performance in PwPD [44], particularly balance [45] and gait performance [46,47]. Otherwise, some results indicate that PwPD can unconsciously couple body sway to visual information to control the postural sway of likely healthy participants [48]. Although previous evidence has demonstrated that KC combined with visual and auditory feedback may enhance motor learning in healthy people [49], PwPD may particularly take advantage of visual and auditory cues offered by games. Moreover, the preserved visual-motor coupling allows the use of visual feedback to improve posture. Further studies are needed to clarify if the addition of KC during the VRG training may be helpful in other populations.

Concerning the KC features, during the Stack Em up game, KC was needed across multiple body regions, including the trunk, elbow, and hips, compared to other games that only required cues for one or two regions. Although various parameters for providing kinesthetic feedback have been studied, the role of variability has yet to be investigated. The high complexity of the game, increased body areas stimulated by KC, or both could explain the observed improvement in the learning process for the Stack Em Up game in the KCG. Further studies should explore the effects of game complexity and KC variability on motor improvement.

The discrete additional gain associated with KC addition was not enough to promote further improvement in functioning. Then, concerning the rehabilitation purposes, the visual and auditory provided by the VRG system were enough to facilitate the improvement in the motor and cognitive function in PwPD. In other words, the addition of KC provided by a PT during the VRG training was not helpful in promoting further functioning improvements. The motor and cognitive features of the participants may have contributed to this finding. Even the participants in the moderate stage of PD (HY 3) could safely keep the standing position to play the games. Then, they could improve their performance regardless of KC provided by PT. Supposedly, the KC could be helpful for training people in the more advanced stage of PD, but further studies should investigate it.

The functioning level is about having the capabilities that enable all people to be and do what they have reason to value [50]. In this study, we adopted functioning as the outcome once this consists of the main rehabilitation goal. The functional status assessment was carried out using the ICF classification. Then, we included several tools related to Function, Activity, and Participation domains. Although a pilot study had shown similar results, we included a wider range of tools related to each ICF domain to offer a more comprehensive evaluation of VRG effects in PD [51]. Our results show the benefits of this kind of intervention for the three ICF domains. This finding reinforces the potential of VRG training for the rehabilitation of people in early to moderate PD stages.

However, we should note that this kind of intervention was not helpful in improving mental function, despite the gains in motor function, Activity, and quality of life. In this study, the exclusion of participants with severe depression may have influenced the results. Considering the high prevalence of depression in PD, besides the VRG intervention, we should consider offering multidisciplinary care to improve the mental health of PwPD [52,53].

The long-distance walking performance also did not improve after the training. Probably, the short duration of the training and low cardiovascular demand did allow endurance improvements. Previous studies (2018) showed endurance improvement after training based on VRG with longer session duration [51,54]. Further studies are needed to investigate the ideal training duration and intensity to promote endurance improvement in PwPD.

Taken together, our results demonstrate that using VRG intervention can enhance the functioning of PwPD, even without direct physical PT assistance during training. This finding suggests that one PT can provide verbal instructions and safety during training to multiple patients, maximizing the PT-to-patient ratio. Although few studies have discussed the cost behind the proposed interventions, reducing cost is an important aspect to be considered for electing more feasible interventions for chronic disease [55].

In addition, it is essential to understand some limitations of using *XBOX 360* with *Kinect®* as the need for a large space to install the device and for the participant to move around; sometimes offers a low level of visual discrimination. More important, it can be challenging for PT to find the proper position during the training to avoid problems with the movement sensor detector.

The results of this study are limited to a narrow group of PwPD in the early to mild stage of disease evolution, able to play the proposed games in terms of motor and cognitive functions. Therefore, the results cannot be generalized to PwPD with more severe motor and cognitive dysfunction. Future research should consider including a larger sample of participants with these features.

Finally, although VRG has often been used to improve specific motor functions such as balance and gait performance, we understand that the challenge for validating this kind of training for rehabilitation is to increase the findings about its efficacy in improving functioning levels. Improvement in functioning levels depends on motor and non-motor enhancements that can change a person’s ability to cope with everyday life. Therefore, we consider all adopted tools equally important to assess it. Further studies should explore identifying which is the primary outcome to enhance functioning level.

## 5. Conclusions

The use of VRG intervention effectively improves functioning in individuals with early to moderate-stage PD, regardless of the addition of KC provided by PT during training.

## Data Availability

All data produced in the present work are contained in the manuscript

